# The combination of glycolic acid and D-lactate delays disease progression in SOD-1 ALS mice, partially rescues lethality in iTBPH^pkk(108354)^ Drosophila and shows promising results in experimental treatments in two ALS patients

**DOI:** 10.1101/2025.11.07.25334423

**Authors:** Alexandra Chovsepian, Yanina Dening, Andrés Jiménez Zúñiga, Carla Palleis, Stefan Sonnenfeld, Guido Rohrer, René Günther, Kai Boetzel, Johannes Levin, Andreas Thalmeier, Jürgen Babl, Peter Falkai, Marianne Dieterich, Adolfo López de Munain, Francisco J. Gil-Bea, Gorka Gereñu Lopetegui, Andreas Hermann, Francisco Pan-Montojo

**Affiliations:** Department of Psychiatry, LMU University Hospital, 80336 Munich, Germany; Neurosciences Area, BioDonostia-BioGipuzkoa Health Research Institute, 20014 Donostia/San Sebastian, Spain; CIBERNED, ISCIII (CIBER, Carlos III Institute, Spanish Ministry of Sciences and Innovation), 28031, Madrid, Spain; Department of Neurology, LMU University Hospital, 81377 Munich, Germany; Department of Neurology, Carl Gustav Carus University Hospital, 01307 Dresden, Germany; German Center for Neurodegenerative Diseases, 81377 Munich, Germany; Hospital Pharmacy, LMU University Hospital, 81377 Munich, Germany; Department of Neurology, Donostialdea Integrated Health Organization, Osakidetza Basque Health Service, 20014 Donostia/San Sebastian, Spain; Department of Neurosciences, Faculty of Medicine and Nursery, University of the Basque Country UPV/EHU, 20014 Donostia/San Sebastian, Spain; Department of Medicine, Faculty of Health Sciences, University of Deusto, 48007 Bilbao, Spain; Department of Health Sciences, Public University of Navarra (UPNA), 31006 Pamplona, Spain; IKERBASQUE, Basque Foundation for Science, 48013 Bilbao, Spain; Department of Physiology, Faculty of Medicine and Nursery, University of the Basque Country UPV/EHU, 48940 Leioa, Spain; Translational Neurodegeneration Section “Albrecht Kossel”, Department of Neurology, University Medical Center Rostock, University of Rostock, Rostock, Germany; German Center for Neurodegenerative Diseases (DZNE) Rostock/Greifswald, Rostock, Germany; Center for Transdisciplinary Neurosciences Rostock (CTNR), University Medical Center Rostock, University of Rostock, Rostock, Germany; Neurological Clinic Sorpesee, Langscheid, Germany

## Abstract

Amyotrophic Lateral Sclerosis is the most common motor neuron disease. It is incurable and, at the time of this study, only two treatments with a limited therapeutical effect are available: riluzole and edavarone (not in Europe). These treatments have been shown to delay the progression of the disease by a maximum of 10%. We recently showed that glycolic acid (GA) and D-lactate (DL) are able to revert some of the phenotypes observed in iPSC-derived neurons from FUS- and SOD-1-ALS patients in vitro. Here we show that the administration of GA and DL is able to delay the progression of the disease in SOD1-G93A mice and protect spinal motor neurons against neuronal death. Interestingly, GA and DL were also able to attenuate the lethality in the TBPH silencing strain (iTBPH^pkk(108354)^) in Drosophila, showing a conserved role between species. Based on these results, we performed two experimental treatments in ALS patients carrying a disease-causing mutation in FUS and SOD-1 respectively. As GA and DL have been shown to be toxic (kidney stones, altered hepatic metabolism) and even lethal above certain doses in humans, we developed and used a specific formulation containing L-alanine to avoid these side effects. Our results show that, together with L-alanine as supportive treatment, GA and DL were well tolerated by the patients. Although promising, well designed and placebo-controlled clinical trials need to be performed in order to confirm the good tolerability and the therapeutic effects in ALS patients.

## Introduction

Amyotrophic Lateral Sclerosis (ALS) is the most common motor neuron disease, with an estimate of 17.000 patients and approximately 5.000 new cases annually only in Europe [1]. Worldwide incidence is approximately 1.6 cases per 100,000 persons annually [2]. Compared to other neurodegenerative disorders, ALS exhibits the fastest fatality rate, with an expected survival time of 2–3 years [2, 3]. Until today, ALS is an incurable disease with Riluzole, Edaravone and Tofersen being the only approved and commercially available treatments (Edaravone only in the USA, Japan and Switzerland, Tofersen only for SOD1 related forms) [4, 5] and a new treatment with the antisense oligonucleotide jacifusen for FUS-ALS [6].

At the time this study was performed, only Riluzole and Edaravone were available for the treatment of ALS. However, the efficacy of these treatments is quite low and the results in real-world settings are mixed [7] [8, 9]. Overall, Riluzole can be expected to delay time to death or time to tracheostomy for patients with ALS by about 10% at best [10] and Edaravone might delay motor deterioration when initiated early [11] but this effect was not reproduced in other studies [8, 9]. All other available treatments, including physical therapy and palliative treatment [12, 13] as well as tracheostomy, chronic mechanical ventilatory support [2], are only symptomatic and do not delay the disease’s progression, with the exception of noninvasive positive pressure ventilation that delays progression in spinal onset patients.

Considering the pathophysiology of ALS, the disease appears in familial (∼10% of cases) and sporadic forms (∼90% of cases) [14] and is caused by the degeneration of motor neurons in the cortex (upper motor neurons) and, the brain stem/spinal cord (lower motor neurons) progressively resulting in paralysis and death [15]. Mutations in the genes C9orf72, SOD-1, FUS, and TARDBP are the most frequent genetic mutations associated with familial ALS in Europe [16]. The pathogenic mechanism underlying motor neuron death has been extensively studied. Increased glutamate signaling and intracellular calcium levels (excitotoxicity), endoplasmic reticulum (ER) stress, mitochondrial dysfunction, oxidative stress due to the increase of reactive oxygen species (ROS), dysregulated transcription and RNA processing, protein misfolding and aggregation, dysregulated endosomal trafficking, impaired axonal transport and neuroinflammation are key components involved in the pathogenesis of ALS as summarized by Tadic et al., 2014. [17]. The particular vulnerability of motor neurons compared to other neuronal groups are still of debate, however might be at least partially explained by their high expression of AMPA receptors that lack the calcium impermeable GluR2 subunit, which makes them more prone to excitotoxicity and imbalances in intracellular Ca^2+^ homeostasis [18]. Moreover, motor neurons are known to express low levels of Ca^2+^ -buffering proteins which increases their vulnerability [19]. Motor neurons strongly rely on optimal mitochondrial function, due to their high metabolic demands and are therefore more prone cell death when mitochondrial activity is dysregulated. Overall, the crosstalk between calcium, ER and mitochondria function as well as oxidative stress seems to be crucial in the development of ALS pathology [17].

In this study, we follow-up our previous study on a novel treatment for ALS based on a combination of two substances: Glycolic acid (GA) and D-Lactate (DL). Here we tested the effect of this combined therapy in Drosophila and mice models of ALS and in a compasive pilot non controlled experimental treatment in two patients carrying mutations in FUS and SOD1 genes respectively. Both compounds occur naturally in the cell as products of DJ-1 [20], a Parkinson’s disease- related enzyme [21], which converts the reactive aldehydes glyoxal and methylglyoxal to GA and DL, respectively [20, 22]. There is accumulating evidence showing that GA and DL have the potential to counteract several of the aforementioned pathological factors. More specifically, both substances can help maintaining calcium homeostasis by reducing calcium influx in the cell [23]. Moreover, in a recent study implementing motor neurons differentiated from induced pluripotent stem cells (iPSCs) derived from fibroblasts of ALS patients with different mutations, we showed that GA and DL together could rescue mitochondrial impairments in FUS- and SOD1-ALS [24]. GA+DL rescued mitochondrial hyper-elongation and restored inner membrane potential in SOD1-ALS axons [24]. In addition, in FUS-ALS axons, GA+DL restored the axonal motility, fragmentation and depolarization of distal mitochondria. GA+DL also restored cytoplasmic mislocalization of FUS and FUS recruitment to DNA damage [24]. This research, in accordance with previous results from our group [25] also suggests that GA can improve the mitochondrial energy production by increasing the levels of NAD(P)H [24]. Finally, it was recently shown that GA can combat oxidative stress via the conversion of glycolic acid into reduced Glutathione [26]. Based on all this evidence, we decided to assess the efficacy and tolerability of GA+DL in delaying ALS progression in 3 different in vivo settings:

1. on the gold standard ALS mouse model (the SOD1-G93A mouse line) via chronic oral gavage administration and compared to the standard of care (Riluzole) or via chronic intrathecal administration.
2. on a Drosophila ALS model silencing muscle-conditioned TBPH Drosophila gene, the fly ortholog gene of human TARDBP.
3. by performing individual experimental treatment with orally administered GA+DL in 2 ALS patients with disease causing mutations in FUS or SOD1, respectively.

In summary, our study is the first to highlight the therapeutic potential of combination treatment with GA+DL in ALS *in vivo.* In the SOD1-G93A mouse model for ALS, oral GA+DL treatment resulted in significant improvements in neuromuscular function and motor neuron survival compared to vehicle and even outperformed Riluzole, the current standard of treatment. Intrathecal administration of GA+DL in this mouse model had also a positive functional outcome compared to vehicle, especially on weeks 15-16 of age, which was also confirmed histologically by the presence of higher numbers of motor neurons in the spinal cord. In drosophila, treatment with GA+DL attenuated the lethality rate of the TBPH silencing strain (iTBPH^pkk(108354)^), showing a conserved effect between different species. Finally, in both ALS patients, treatment with GA &DL using a specific formulation was well tolerated without causing renal or liver damage and seem to have a positive effect on disease progression. Creatinine Kinase levels were reduced upon treatment, with the most rapid decline evident in the SOD1 patient.

## Materials and Methods

### Animals, experimental design and timeline

#### SOD-1 Mouse model

All animal procedures complied to the German guidelines on animal welfare and were approved by local regulatory committees (Regierung von Oberbayern, ROB-55.2-2532.Vet_02-19-149).

46 transgenic hemizygous males expressing a high copy number of the human SOD1 gene with a G93A mutation (B6SJL-Tg(SOD1*G93A)1Gur/J) were obtained from the Jackson Laboratory at an age of 7 weeks. 21 of them received oral treatment (GA+DL/ Riluzole/ Vehicle; n=7) and 25 of them received treatment intrathecally (GA+DL; n=14 or Vehicle; n=11). 25 WT male mice (C57BL/6; Janvier Labs, France) of the same age were used as non-transgenic controls (n=12 for oral and n=13 for intrathecal administration). After a week of habituation, we started the Neuroscore evaluation (8-weeks old mice) twice per week. Oral treatment (GA+DL/ Riluzole/ Vehicle, once per day, 6 days per week) or intrathecal administration started at the end of week 10 for the next 6 weeks. Mice were perfused by the end of week 17 for histological analysis.

A schematic of the experimental timeline and number of mice per treatment group is provided in Fig. 1.

**Figure 1:**
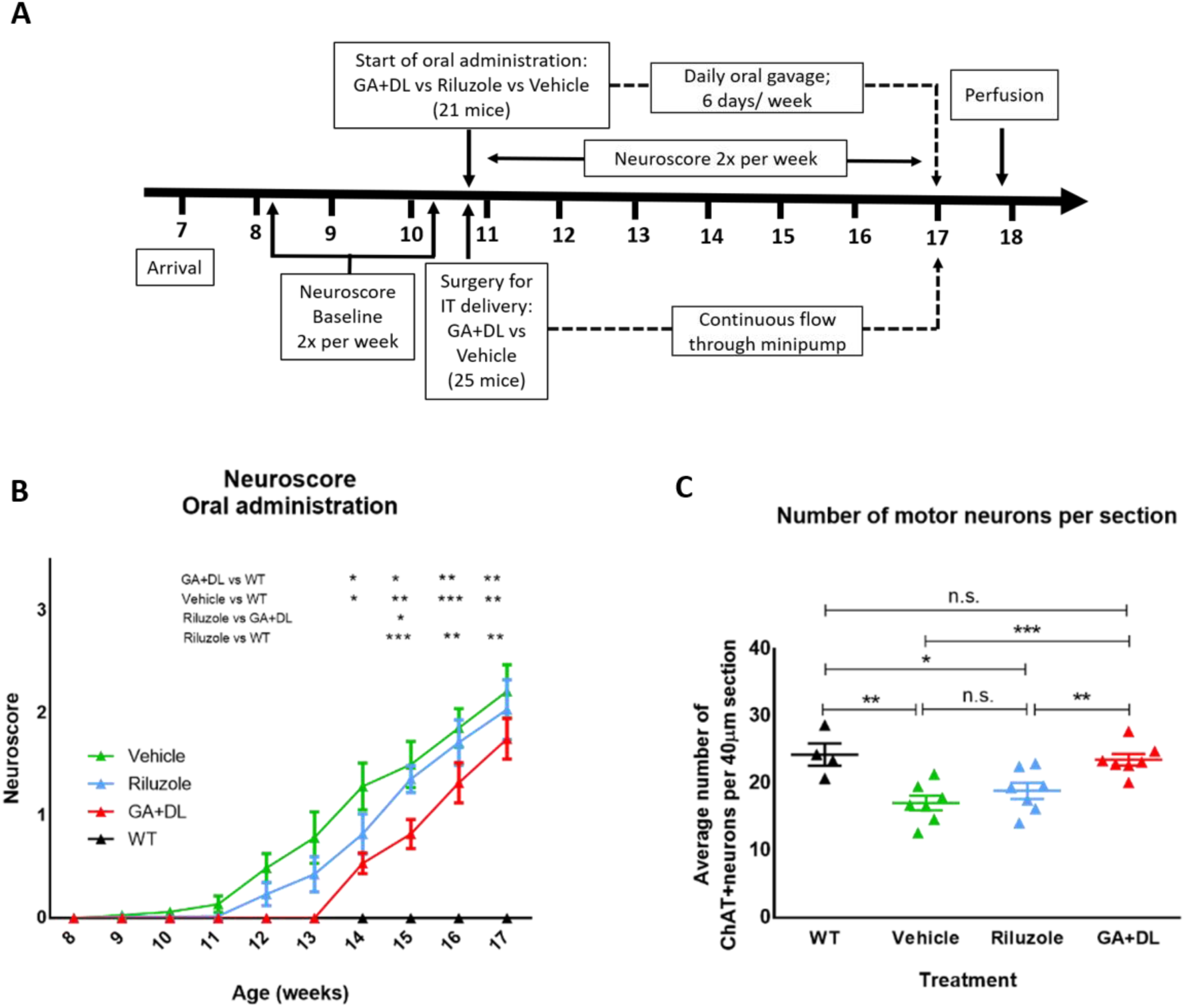
A: Experimental timeline: 46 transgenic hemizygous males expressing a high copy number of the human SOD1 gene with a G93A mutation (B6SJL-Tg(SOD1*G93A)1Gur/J) were obtained from the Jackson Laboratory at an age of 7 weeks. 21 of them received oral treatment (GA+DL/ Riluzole/ Vehicle; n=7) and 25 of them received treatment intrathecally (GA+DL; n=14 or Vehicle; n=11). 25 WT male mice (C57BL/6; Janvier Labs, France) of the same age were used as non-transgenic controls (n=12 for oral and n=13 for intrathecal administration). After a week of habituation, the Neuroscore evaluation (8-weeks old mice) took place twice per week. Surgical implantation of catheter and minipump for intrathecal administration started at the end of week 10, and the pumps run continuously for the next 6 weeks. The daily oral gavage treatment (GA+DL/ Riluzole/ Vehicle, once per day, 6 days per week) started at the end of week 10 and lasted for the next 6 weeks. Mice were perfused by the end of week 17 for histological analysis. B, C)Graphs showing the functional (B) and histological (C) outcome of orally administered GA+DL in the SOD1-G93A mouse model for ALS. B): Neuroscore values: 0= pre-symptomatic; 0= symptom appearance; 2= paresis onset; 3= paralysis; 4= humane end-point. 2-way ANOVA tests showed that each of the SOD1 treatment groups had worse functional outcome than WT (p<0,0001). However, GA+DL treatment significantly improved neuromuscular function compared to vehicle (2-way ANOVA, p=0,014) and compared to riluzole treatment (2-way ANOVA, p=0,013). The Neuroscore upon riluzole treatment did not differ from vehicle treatment (2-way ANOVA, p=0,24). Stars signify post-hoc significant differences at the corresponding timepoints. C): Average number of ChAT^+^ motor neurons in the ventral horn of the lumbar spinal cord of wild-type (WT), or SOD-1 (Vehicle, riluzole, GA+DL) mice treated orally. A total of 14 sections (40 µm) per mouse separated by 240 µm (1:6 section) were quantified and the average amount of motor neurons per section was calculated. Vehicle (water) treated SOD-1 mice showed a significant loss of motor neurons in this region when compared to WT. While oral treatment with riluzole (the standard-of-care in Europe) did not protect against this loss, treatment with GA+DL rescued this phenotype and protected against neuronal loss.

### Preparation of solutions for oral administration

#### GA+DL solutions

GA+DL stock solution (3.819M in sterile ddH2O, pH adjusted to 7.48 with NaOH 15M) was prepared under sterile conditions. The solution had to be vortexed and sonicated, always kept on ice during the process. GA+DL solution for oral administration (100mg/kg) was obtained by diluting the aforementioned stock solution in H2O to achieve the target concentration of 0.26 M and stored in aliquots at -20°C, to be used fresh, daily.

#### Riluzole solution

Riluzole solution was prepared in accordance with Hogg et al., 2018 [27]. A Rilutek tablet (50mg) was dissolved in 2.27 ml DMSO to acquire a stock solution of 22mg/ml. The solution had to be vortexed and sonicated, always kept in ice during the process. The stock solution was further diluted in H2O to the target concentration of 1.6mg/ml (8 mg/kg) and stored in aliquots at -20°C, to be used fresh, daily. Before use, each new aliquot was again sonicated to ensure a homogeneous delivery.

#### Vehicle

For the oral administration experiments H2O from each mouse cage’s drinking bottle was used as Vehicle.

### Preparation of solutions for intrathecal administration

The target CSF concentration of GA+DL to be achieved in the CSF is 10mM. For that purpose, the calculated minipump reservoir concentrations are 152-fold higher (CSF production: 22.8 µL/h; pump rate: 0.15 µL/h). Therefore, the concentration of GA+DL in the minipump solution was 1.52M. Both substances were dissolved in ddH2O under sterile conditions and the pH was adjusted to 7.33 with NaOH. In the vehicle administration group minipumps were filled with 0.9% NaCl. The preparation of solutions and filling of the minipumps was carried out in sterile conditions.

### Minipump implantation for intrathecal delivery

Before starting the implantation process, mice were given peroral analgesia (5 mg/kg meloxicam) and then injected i.p. with combination anaesthesia (midazolam 5.0 mg/kg + medetomidine 0.5 mg/kg + fentanyl 0.05 mg/kg). To protect the cornea, an eye ointment (Bepanthen) was applied. The body temperature was kept constant via a feedback heating mat. The surgical procedure was followed as previously described by Ineichen et al. [28] with slight modifications. After verifying the lack of pedal reflex, the head was fixed on the stereotactic frame by inserting the ear bars in the ear canals. A rostro–caudal skin incision was performed over the cervical spinal column and occipital bone, and the uppermost muscle layer (trapezius muscle) was split with scissors. The head position was then adjusted so that the nose pointed down with a 90° angle toward the body axis using the mouthpiece for stabilization. The second muscle layer was then pulled apart using forceps. The third muscle layer was disconnected from the occipital bone until the cisterna magna appeared covered by the atlanto-occipital membrane (AOM). Following a careful superficial medio-lateral incision to the AOM and the underlying dura, the brain stem was exposed. The osmotic mini-pump (ALZET Pump Model 2006, #0007223) connected to the intrathecal catheter (Alzet mouse intrathecal catheter, #0007743) filled with the solution of interest was placed on a sterile surface next to the animal. The catheter wasinserted into the intrathecal space in the caudal direction up to 0.8 cm and is then fixed with a droplet of tissue glue and a ligature around the second muscle layer. After suturing the uppermost muscle layer, the pump is placed into the subcutaneous pouch. The catheter is further fixed to the muscle tissue using tissue glue, the wound is closed with sutures and local anesthesia ointment is applied. At the end of the operation, which takes around 20 - 30 minutes per animal anesthesia was antagonized (atipamezole 2.5 mg/kg + flumazenil 0.5 mg/kg + naloxone 1.2 mg/kg, s.c.). The mice woke in a cage on a hot plate (38°C) and received for a further 2 days 5 mg/kg meloxicam orally every 12 hours. The mortality rate during to complications during the operation or just after the operation was 35,7% for all groups.

### Neuroscore test

In order to evaluate the neuromuscular function of our mice, we implemented an established phenotypic protocol (namely ‘NeuroScore’), previously used for SOD1-G93A mice [29]. The NeuroScore test was performed twice per week in the morning and the performance was always recorded on video. The test starts by suspending the mouse by its tail above the mouse cage for 3 sec for observation of its hindlimbs (HL), with a total of 3 repetitions. Then, the mouse is placed on a clean surface with some friction where it is allowed to walk uninterrupted, for careful gait observation. At a later stage of the disease, when paresis has started the mouse is placed on its side (left and right side to be tested) and the time it needs to return to its normal position (righting reflex) is measured. Each HL is scored separately, from 0 (pre-symptomatic) to 4 (rigid paralysis). NS of 0 signifies normal HL splay (full extension away from midline) for 2 sec or more and normal gait, comparable to a healthy naive control. NS of 1 (symptom appearance) signifies an abnormal splay, such as (partial) collapse of the hindlimb towards the lateral midline, retraction or trembling during the tail suspension test. NS of 1 is still given when mouse has a normal or slightly slow gait. A NS of 2 (paresis onset) signifies partial or complete HL collapse towards the lateral midline and evidently reduced extension ability, but not complete lack of movement during tail suspension. For a NS of 2, during gait observation the mouse is using the hindlimb for moving forward but the toes curl downwards or the mouse is dragging the HL. In terms of the righting reflex test, for a NS of 2 the mouse is able to right itself in 10 sec from both sides. A NS of 3 (paralysis) is assigned when, during tail suspension, there is lack of HL movement or minimal joint movement. During gait observation, there is forward motion but without hindlimb use and the righting reflex for both sides does not extend 10 sec. A NS of 4 (Humane end-point) is applies to the following observations: There is rigid HL paralysis during tail suspension, no forward motion when the mouse is allowed to walk and the righting reflex takes longer than 10 sec, at least from one of the two sides.

### Histology

Mice were anesthetized via an isoflurane inhalation mask (2% isoflurane) and i.p. injection of MMF (Midazolam 5,0 mg/kg + Medetomidin 0,5 mg/kg + Fentanyl 0,05 mg/kg). After confirming the absence of posterior interphalangeal and lid closure reflexes, mice were transcardially perfused with 4% paraformaldehyde (PFA) in PBS. The spinal columns were kept in PFA overnight at 4°C and then temporarily stored in PBS until they were micro-dissected under the microscope for spinal cord isolation. The spinal cords were then kept in 50% sucrose solution for 48h and then transferred to 30% sucrose solution for another 48h. Following that, the tissue was embedded in Tissue-Tek® O.C.T.™ for cryoprotection and stored in -80°C. A 10-mm block of lumbar spinal cord was sectioned in 40-µm thick sections with a cryostat (Leica CM1860), and placed in 96 well-plates with freezing medium (25% Glycerol [Sigma], 25% Ethylene Glycol [Sigma], 50% PBS) to be kept in -80°C until immunohistochemical analysis.

### Immunohistochemistry

1 every 6 lumbar spinal cord sections were used for immunohistochemistry. The sections were transferred to 12-well plates and washed with PBS 3x 10min. The sections were then incubated in blocking buffer solution (5% donkey serum, 1%Triton X-100 in PBS) for 2h at room temperature and overnight at 4°C. The next day, the primary antibody (Ab’) solution was applied: Goat anti-ChAT Ab’ (AB144P Sigma-Aldrich) 1:300, 1% donkey serum, 1%Triton X-100 in PBS. The sections remained in the Ab’ solution at room temperature for 2h and overnight at 4°C. Following that, after 3x 10 min washing with PBS, the sections were incubated with the secondary antibody (Ab’’) solution: Donkey anti-Goat-Alexa 555 (A-21432 Invitrogen) 1:300, 1% donkey serum, 1%Triton X-100 in PBS at room temperature for 3 hours. In the last of the following 3x 10min PBS washings, DAPI stain (5087410001 Merck) was added to the PBS (1:1000). The sections were washed one more time with PBS before being mounted on glass slides and covered with coverslips using mounting medium

### Quantification of ChAT+ motor neurons

Lower motor neurons (LMNs) in the ventral horn were identified based on the expression of the molecular marker ChAT and co-presence of DAPI. 10× Images were captured using a Zeiss Apotome 2 microscope. DAPI+/ChAT+ LMNs were quantified for each stained section using Image J (National Institutes of Health, Bethesda, MD) and the average number of LMNs per section was calculated per mouse.

### *Drosophila melanogaster* model and adult pharate survival assay

The *Drosophila* strains employed for these experiments were: *UAS-Dicer2-iTBPHp(pkk108354)* (nRef: #104401 from Vienna Drosophila Stock center (VDRC)) and Dicer2-*Mef2-GAL4 from Bloomington Stock Center (BDSC, nRef:* #25756), for muscle-conditioned silencing. To perform crosses, 10 Virgin *iTBPHp(pkk108354*) females with *3-4 Mef2-GAL4* males were placed each tube and *UAS-Dicer2-iTBPHp(pkk108354)-Mef2-GAL4*, (iTBPH^pkk108354^) flies were finally generated. As control flies, the strain *UAS-Dicer2-+-Mef2-GAL4* was used. As control flies, the strain *UAS-Dicer2-+-Mef2-GAL4* was used Flies were housed at 23°C, 70% humidity and 12 h/12 h light/darkness cycle. Adult pharate survival was expressed as a percentage of the adult flies counted over the total number of pupae in each tube where adult pharate survival of control flies was around 100%. 12 tubes were counted per group in i*TBPH^pkk(108354)^* flies.

### Experimental treatments in ALS patients

In Germany, a medical doctor has the freedom to treat patients with incurable diseases leading to death for which there are no effective treatments in an “individual experimental attempt of healing” or in german “Individueller Heilversuch” and prescribe a medication that is suitable for this disease although it has not yet been tested and is not approved by the Federal Institute for Medicines (BfArM), Paul-Ehrlich-Institut or EMA. ALS qualifies as such a disease. All proceedings described below were presented at the ethic committees of the University Hospital of the LMU (Reg. Nr.: 22-0366UE) and the Carl Gustav Carus University Hospital (Reg. Nr.: EK49022016) who showed no concerns regarding the design of the experimental treatment and publication of the results. Glycolic acid, D-Lactic acid and L-Alanine Ph.Eur. were obtained by the LMU hospital pharmacy. The quality control of Glycolic acid and D-Lactic acid was performed using the methods described in the Deutscher Arzneimittelcodex (German Drug Codex/NRF), L-Alanine Ph.Eur. was tested using the corresponding European Pharmacopoeia monograph.

Two ALS patients were treated using a combination of GA and DL together with L-Alanine. One of the patient carried a FUS mutation (c.1562G>A (p.Arg521His) and the other a heterozygous p.Arg116Gly SOD-1 mutation. The FUS patient was a male in his 40’s. The diagnosis of ALS had been made one year before. At the time of recruitment the strength in the lower limbs was strongly reduced (see evolution of lower limbs strength in Supplementary Figure S2 F and G) and was starting to decline in the upper limbs (see Supplementary Figure S2 A-E). He was under treatment with Riluzole 50 mg twice a day. The SOD-1 patient wasfemale in her 70’s. The diagnosis of ALS had been made one year before recruitment debutating as a leg accentuated tetraparesis (see Table 1 for demographic and clinical observations).

**Table 1.** Demographic, main clinical observations and adverse reactions/events in ALS patients treated with GA & DL

|  | **P 1 = FUS-ALS** | **P 2 = SOD1-ALS** |
| --- | --- | --- |
| **Gender** | male | female |
| **Age at treatment initiation (Y)** | 40 to 50 | 70-80 |
| **Treatment details** | Natriumlactate 13,8ml and Natriumglycolate 13,8ml 0-0-1 | Natriumlactate 13,8ml and Natriumglycolate 13,8ml 0-0-1 |
| **Treatment initiation** | 02.2017 | 07.2017 |
| **Date of high dose reached** | 04.2017 | 08.2017 |
| **End of treatment** | 09.2018 | 02.2018 |
| **Treatment duration (month)** | 17 | 7 |
| **Age at onset (Y)** | 40's | 70's |
| **-- symptoms** | 09/2015 | 05/2015 |
| **-- diagnosis** | 09/2016 | 04/2016 |
| **Genetics** | FUS c.1562G>A (p.Arg521His | SOD1 c.346>G; p.Arg116Gly heterozygote |
| **Family history for ALS** | no | Family member died due to muscle wasting with respiratory insufficiency |
| **Family history for FTD** | no | no |
| **ALS onset type** | spinal | spinal |
| **ALSFRSR at treatment onset** | 35 | 34 |
| **Clinical presentation to date of treatment onset** | Paraparesis (spasticity?) without signs of bulbar involvement. | Leg-accentuated tetraparesis without spasticity and no signs of bulbar involvement |
| **Laboratory findings** | Elevated levels of: CK, AST and ALT. CK was already increased before treatment and decreased with treatment. ASL and ALT increases were limited in time and due to alcohol | Initially slightly increased Troponin T without increase during treatment |
|  | consumption. Transitory increase in oxalate crystals in the urin, reduced after the increase of the L-Alanine dose. | Creatinine was already decreased before treatment initiation and did not increase during treatment, creatinine-GFR was normal<br>AST, ALT, gGT, LDH, GFR normal<br>CK was slightly increased before treatment initiation und decreased during treatment |
| **Time on GA &DL (weeks)** | 72 weeks | 38 weeks |
| **Co-medication** | Riluzole 50mg 1-0-1<br>Amlodipin 5mg 1-0-0<br>Lorazepam 1mg bB | Riluzole 50mg 1-0-1<br>Tilidin 50/4mg 1-0-1<br>Novaminsulfon 40Trpf. 1-0-1 und b.Bed.<br>Imatinib 400mg Mo-Fr und WE Pause<br>Alendronsäure 1x/Wo<br>Calcilac 1x tgl.<br>Enoxaparin 40mg 1-0-0<br>Pantoprazol 40mg 1-0-0 |
| **Adverse reactions or events** | No treatment related adverse reactions or events | No treatment related adverse reactions or events |

ALS: amyotrophic lateral sclerosis; PRN – pro re nata; Y – years; AST: aspartate transaminase; ALT: alanine transaminase; CK: creatine kinase; LDH: lactate dehydrogen

The treatment took place once a day for 6 days a week. The toxicity of GA and DL has been already tested on animals. The reported LD50 (lethal dosage for 50% of the population) of GA for rats and pigs was 1950 and 1920 mg/kg body weight, respectively. However, other report in rats treated with glycolic acid at 0.6% of body weight no side effects were observed [30]. Glycolic acid was also tested in cats, a dose of up to 100 mg/kg body weight had no side effects. One daily intake (acceptable daily intake (ADI)) up to 100 mg/kg body weight of D-lactic acid has already been approved by the American FDA [31]. Based on our results in mice and the above-mentioned toxicity information, we started with low doses of the compounds and increased them weekly. Laboratory controls were performed once a week until the target concentration was reached and then once every two weeks. The dose increase was performed as described in Table 2:

**Table 2:**
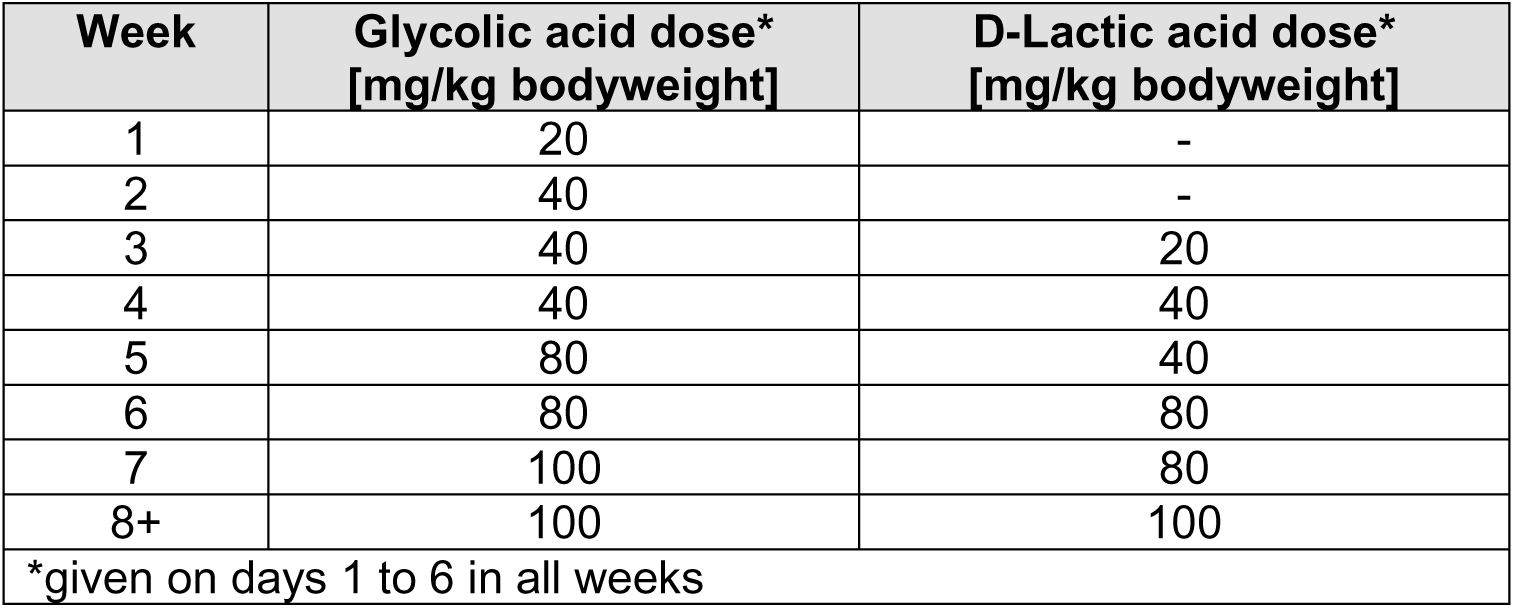
GA and DL dosing

Additionally L-Alanine was also given with increasing doses until reaching a maintenance dose of 3 gramm per day. Both patients were additionally treated with riluzole for the whole duration of the experimental treatment, the standard of care treatment in Europe.

## Results

### Orally administered GA+DL delay disease progression in the SOD1G93A mouse model of ALS

First, we decided to test the effects of chronic oral (via gavage) administration of GA+DL *in vivo*, using the well-studied SOD1- G39A ALS genetic mouse model which is still the gold standard in ALS preclinical research. In terms of the functional impairment induced by the mutation in the SOD1- G39A mouse model, mild motor symptoms can be observed at about 7-8 weeks of age [29]. At approximately 110-120 days of age, motor symptoms become more apparent, such as tremor in at least one limb which progressively affects all limbs [32]. Muscle weakness, muscle atrophy and neuromuscular junction denervation advance, ultimately resulting in paralysis and death [33]. SOD1- G39A mice also exhibit some of the histological hallmarks in ALS such as loss of lower and upper motor neurons. The pathology is more prominent in the spinal cord (predominantly in the lumbar part) and neuromuscular junctions.

We compared the effect of 6-week daily oral gavage administration of 100 mg/kg GA+DL to vehicle (H2O) and to the standard of care (Riluzole) at a concentration of 8 mg/kg which has been demonstrated to be effective in delaying the muscle strength loss in mice with progressive motor neuropathy [34]. The efficacy of each chronic treatment was evaluated in terms of survival, neuromuscular function and motor neuron cell death of our female SOD1-G93A vs female WT (C57BL6/J) mice of the same age. As shown in our experimental timeline graphic (Figure 1 left) the evaluation of neuromuscular function was initiated at the age of 8-weeks using a previously established paradigm (‘Neuroscore’ [29]) and performed twice per week until the end of the experiment. The oral administration of GA+DL/riluzole/vehicle (water) started at the end of week 10 and was performed once daily, 6 days per week. All mice were perfused by the end of week 17 for histological analysis, to quantify the surviving motor neurons in the lumbar spinal cord. For this purpose, mice were transcardially perfused at the end of week 17 (or earlier if they met the humane endpoint criteria). The fixated tissue was processed and stained for identification of surviving motor neurons (ChAT immunofluorescence) in the ventral horn of the lumbar spinal cord. In this model, treatment with the GA+DL combination resulted in significant improvements in the neuromuscular function (Figure 1, middle). All SOD1 groups showed significant functional deterioration compared to WT (2-way ANOVA, p<0.0001), independent of the treatment. The clinical phenotype became apparent between week 13-14. However, 100mg/kg GA+DL treatment resulted in improved scores compared to vehicle (2-way ANOVA: p= 0.014; Sidak’s multiple comparisons test: Timepoints 13, 14, 15 **) and riluzole (2-way ANOVA p=0.013, respectively. Sidak’s multiple comparisons test: Timepoint 15 **). The riluzole-treated group performance did not significantly differ from vehicle (2-way ANOVA p=0.24). Histologically, as can be observed in Figure 1 (right), oral treatment with GA+DL significantly improved the loss of motor neurons in the lumbar spinal cord (1-way ANOVA, p=0.007, followed by Tukey’s post-hoc test), when compared to vehicle (p=0.0003 or riluzole group (p=0.0046). WT mice had significantly higher numbers of motor neurons compared to the vehicle group (p=0.0022) as well as the riluzole group (p= 0.0134). On the contrary, GA+DL treated mice showed an amount of motor neuron in the anterior horn of the lumbar spinal cord not significantly different from WT mice (p=0.336), therefore highlighting the superiority of GA+DL treatment compared to the standard of treatment.

### Intrathecal administration of GA+DL increases the effect of these substances on the progression of motor symptoms in SOD1G93A mice

We followed the same experimental timeline as the one shown in Figure 1, with intrathecal administration starting at the end of week 10. The chronic IT administration is achieved using a delivery method that has been standardized in rodents [28] and involves the subcutaneous implantation of an osmotic minipump loaded with a GA+DL solution that would achieve 10 mM GA+DL in the CSF, or loaded with vehicle (NaCl). The minipump is connected to a catheter which is carefully inserted into the subarachnoid space through an opening in the *Cisterna magna*. The administration is continuous and can last up to 5-6 weeks according to the minipump manufacturer (Alzet, USA). As this method has been shown to not induce any major histological damage, immune reaction or glial activation [28], it is considered safe and is expected to achieve direct delivery of GA and DL to the CNS with minimal systemic exposure. The overall mortality of the procedure was 35,7%, mostly due to intraoperational complications. The effects of IT administration were assessed using the same functional and histological tests as the ones performed in the oral administration experiments.

Our results (Figure 2, left) showed that while vehicle-treated SOD1 mice were performing significantly worse than WT (2-way ANOVA, p<0.0001), GA+DL-treatment resulted in functional outcomes that were not significantly different to WT (2-way ANOVA, p= 0.34). Furthermore, GA+DL improved the neuromuscular function compared to vehicle, especially on week 15 and 16 (2-way ANOVA, p=0.028 followed by Sidak’s multiple comparisons). Histologically, IT-administered GA+DL reduced the loss of motor neurons in the lumbar spinal cord compared to Vehicle treatment. In addition, the GA+DL treated group did not significantly differ from control (Figure 2, right).

**Figure 2:**
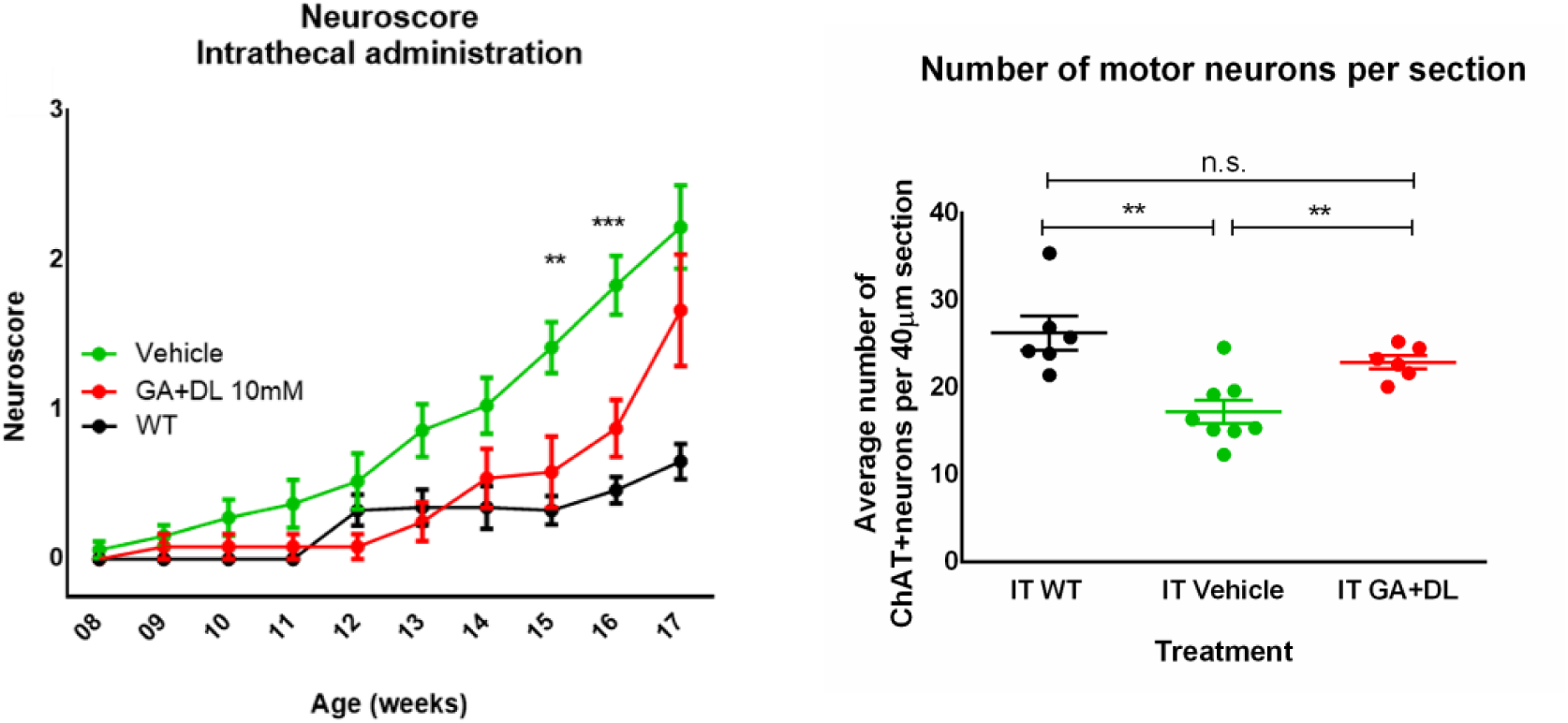
Graphics showing the functional (left) and histological (right) outcome of orally administered GA+DL in the SOD1-G93A mouse model for ALS. **Left:** Neuroscore evolution from week 8 to week 17 of age in the different treatment groups. Vehicle scores were significantly worse than GA+DL (2-way ANOVA, p=0,028). 2-way ANOVA test showed significant deterioration in Vehicle vs WT (p<0,0001) while the functional outcome between GA+DL vs WT was not significantly different (2-way ANOVA, p= 0.34). Interestingly, the moment the pumps were empty according to the manufacturer specifications (end of week 16), there was a clear worsening of the Neuroscore. Stars signify post-hoc significant differences between GA+DL and Vehicle at the corresponding timepoints. **Right:** average number of ChAT^+^ motor neurons in the ventral horn of the lumbar spinal cord of wild-type (WT), or SOD-1 (Vehicle, GA+DL 10mM) mice treated intrathecally (IT). A total of 14 sections (40 µm) per mouse separated by 240 µm (1:6 section) were quantified and the average amount of motor neurons per section was calculated. Vehicle (NaCl 0.9%)-treated SOD-1 mice show a significant loss of motor neurons in this region when compared to WT (ttest IT WT vs IT Vehicle: p=0.001) but treatment with GA+DL rescued this phenotype and protected against neuronal loss (ttest GA vs IT Vehicle: p=0.0059; IT WT vs IT GA: p= 0.145).

### GA & DL attenuate adult pharate lethality rate in a Drosophila model of ALS

Having shown a positive effect on neuronal survival and functional outcome in an ALS mouse model, our next step was to investigate whether this effect is conserved across species. To that end, we aimed to assess the effects of GA+DL treatments on muscle function *in vivo*, generating a *Drosophila melanogaster* model with conditional silencing of *TBPH* gene (human *TARDBP* ortholog) by dsRNA specifically in muscle tissue (Fig. 3a). *TBPH* knockdown were conditioned to the expression of the Myocyte enhancer factor 2 (*DmeI/Mef2*) gene, a transcription factor essential for myogenesis [35, 36]. This Drosophila model shows a significant decrease in *TBPH* gene expression [37].

**Figure 3.**
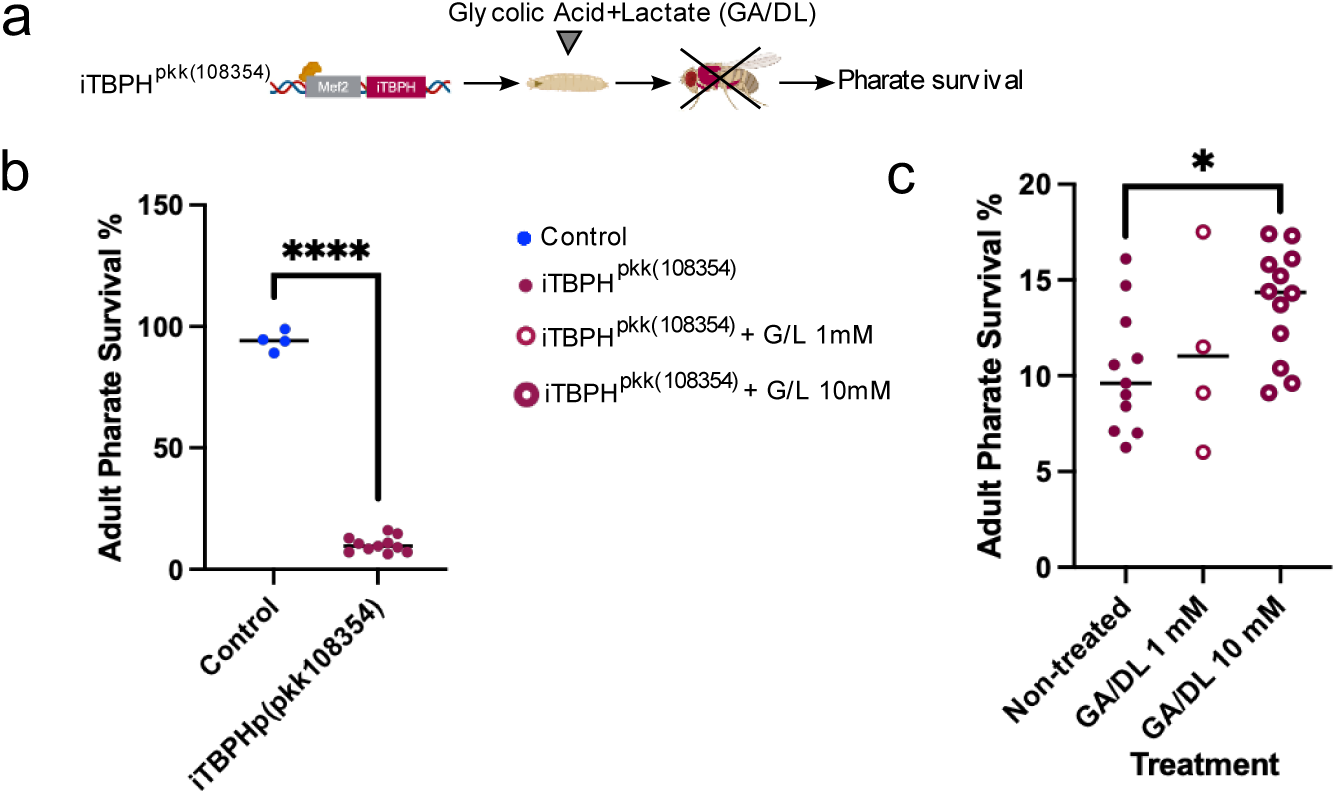
Glycolic acid/D-Lactate treatments partially revert adult pharate survival functional parameter in a *Drosophila melanogaster* model of ALS. A) Schematic illustration of the TBPH silencing fly model (*iTBPH^pkk(108354)^*), the ortholog gene of human TARDBP, used in this study and the analyses performed. Treatments has been administrated orally (by feeding). B) Scatter dot-plots with median and interquartile range showing a dramatical impairment in eclosion efficiency rate induced by muscle-conditioned TBPH silencing. Note that the adult pharate survival of non-silenced flies is close to 100%. However, adult pharates of the *iTBPH^pkk(108354)^*model remain trapped inside the pupa. \*\*\*\**p*<0.0001 compared to control flies via Student’s t test. C) Effect of Glycolic acid/D-lactate treatments (1mM and 10mM) on adult pharate survival in muscle-conditioned TBPH silencing flies. 3 different crosses per group with four different stages for each cross (non-treated and 10mM treatment); 4 different crosses for 1mM treatment. \**p*<0.05 One-way ANOVA followed by Bonferroni post-hoc comparisons.

The TBPH silencing strain (iTBPH^pkk(108354)^) exhibited lethality in newly metamorphosed adults (pharate), not even being able to hatch out of its pupa (Fig. 3b, p<0.-0001) while physiologically near to 100% of flies are able to complete its development, therefore this assay is suitable to test the effect of Glycolic +Lactate (G/L) treatment for ALS.

We applied GA/DL treatment in *iTBPH* flies at two different concentrations. While GA/DL treatment at 1mM had not exert an improvement in adult pharate survival outcome (ns), when we used a higher concentration, 10mM of GA/DL treatment, significantly improved adult pharate survival of these flies (Figure 3c, p<0.05), possibly by enhancing muscle strength to hatch out as suggested by Zufiria M and cols [37]. This was evidenced by a substantial increase in survival rate, from 9% to 14%, at the pharate pupal stage (Fig. 3c).

### GA & DL within a specific formulation containing L-alanine is safe in two individual experimental treatments in familial ALS patients

After showing that the combination of GA and DL had a very strong positive effect in mice and flies, we asked ourselves, whether a similar treatment would be possible in humans. The combination GA & DL has not yet been tested as a drug in humans and the studies on GA ingestion from the 80s and 90s were not very promising. GA was thought to be the main toxic agent in patients that had consumed Austrian wine adulterated with diethylene glycol during the wine scandal of 1985 [38]. D-lactate was considered the key player in the development of encephalopathy in short-bowel syndrome [39]. We finally asked ourselves whether the intake of GA & DL is safe in amounts delivering enough CNS concentration compared to our *in vitro* and preclinical *in vivo* model systems.

Since our previous study showed responsiveness of iPSC-derived motor neurons specifically of FUS- and SOD1-ALS patients, one patient of each gene received an experimental oral treatment with GA & DL (up to 100mg/kg daily) and L-alanine (up to 3 grams/day) (for details see Table 1 and treatment protocol in M&M). The SOD1 patient was afemale in her 70’s with a positive family history carrying a heterocygous p.Arg116Gly SOD1 mutation. She had spinal onset disease; leg accentuated atrophic tetraparesis with ALSFRSR score at begin of treatment of 34. Treatment duration was seven months and complicated by a broken leg which was unrelated to the treatment. There was no reported adverse event, clinical course is depicted in Figure S1.

The FUS patient was amale in his 40’s. The diagnosis of ALS had been made one year before. At the time of recruitment, the strength in the lower limbs was strongly reduced (see evolution of lower limbs strength in Supplementary Figure S2 F and G) and was starting to decline in the upper limbs (see Supplementary Figure S2 A-E). The duration of the experimental treatment in the FUS patient was 1 year and 6 months. The treatment was well tolerated by the patient for the whole treatment time. Apart from a transitory increase in oxalate crystals in the urine that disappeared after increasing the dose of L-alanine, no signs of hepatic or renal toxicity were observed (see Figure 4D-F). The patient initially reported a subjective mild improvement of the strength in the upper limbs, that was objectified by the gripping force measurements (see Supplementary Figure S2E), in general the progression of the strength loss seemed to be slower than within the lower limbs. This was associated with a reduction in CK blood levels see Figure 4. Individual clinical course during treatment can be seen in Supplementary Fig 2 and 3 (SOD1 and FUS in separate Suppl. Figures). The treatment ended when a complete tetraparesis set in and the patient had problems with swallowing the medication.

**Figure 4:**
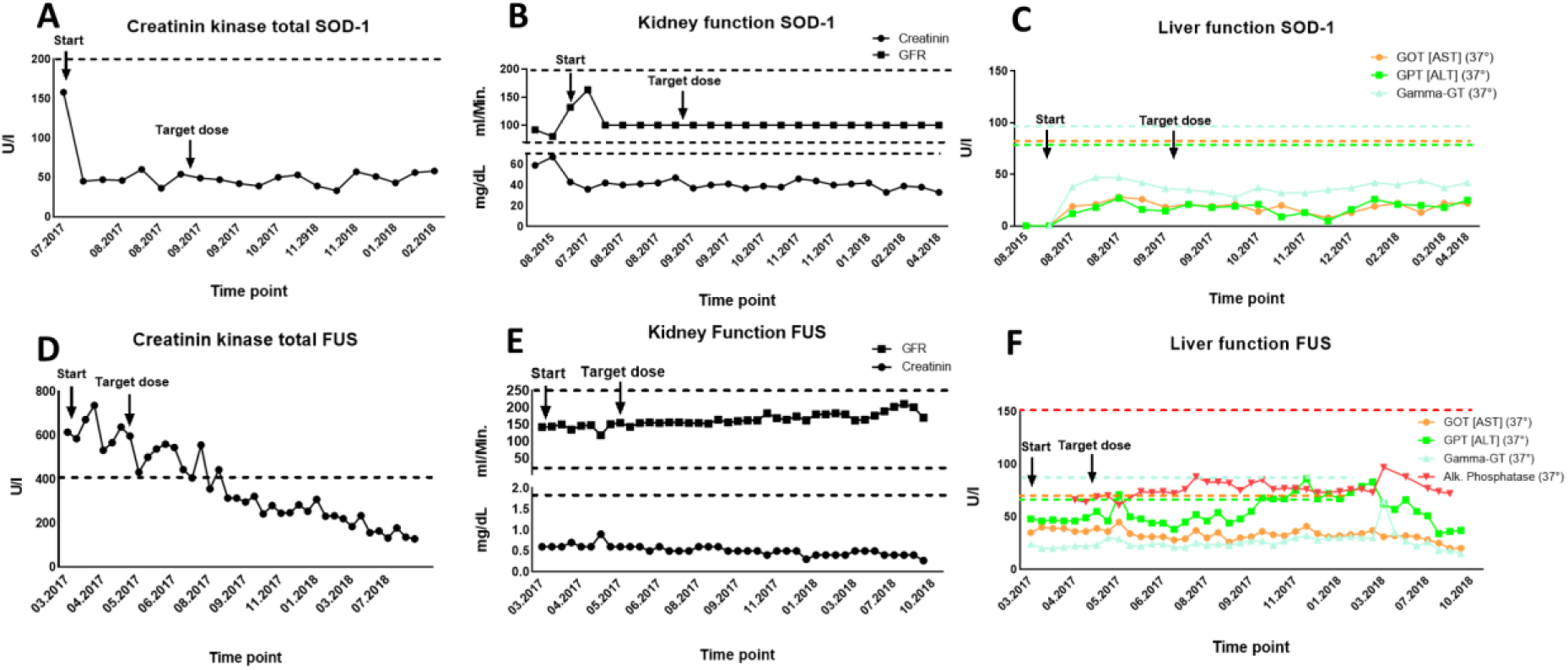
Clinical values of ALS patients with both mutations. **A)** Time course of Creatinine Kinase levels in the blood of the SOD-1. **Β)** Time course of Creatinine and glomerular filtration rate (GFR) levels in the SOD-1, patient indicative of normal kidney function. **C)** Time course of aspartate aminotransferase (AST or GOT), alanine transaminase (ALT or GPT) and gamma-glutamyl transferase (GGT) levels in the SOD-1patient, showing no signs of liver dysfunction. **D)** Time course of Creatinine Kinase levels in the blood of the FUS patient. **E)** Time course of Creatinine and glomerular filtration rate (GFR) levels in the FUS patient. **F)** Time course of aspartate aminotransferase (AST or GOT), alanine transaminase (ALT or GPT) and gamma-glutamyl transferase (GGT) levels in the FUS patient. The transitory increase in GPT levels was due to time-limited alcohol ingestion.

## Discussion

In this study, we validated a putative novel treatment for ALS based on a combination of the two substances glycolic acid (GA) and D-Lactate (DL) *in vivo*. Both compounds occur naturally in the cell as products of DJ-1 [20], which converts the reactive aldehydes glyoxal and methylglyoxal to GA and DL, respectively [20, 22]. We show that GA & DL delayed disease onset and reduced loss of spinal motor neurons in SOD1-G93A ALS mice, in both oral and intrathecal delivery. Interestingly, mice treated with the intrathecal delivery of the substances showed a stronger treatment effect than oral treatment and a rapid worsening of the symptoms once the osmotic pumps were empty, which in a way mimics the design of a cross over study. This could be an interesting approach for the design of a clinical trial in human similar to the design of Nusinersen for SMA therapy.

Although the number of surviving motor neurons in the spinal cord measured on week 17 is higher in the GA+DL treated groups, the function had already been similarly lost at week 17 in all transgenic groups. Previous studies have shown that axonal degeneration with functional consequences can precede motor neuron death in the spinal cord [40]. Therefore, it has to be assumed that in GA+DL treated mice an axonal degeneration had already started, even if the soma of the motor neurons in the spinal cord were preserved, which would explain the delayed appearance of the motor symptoms and further confirm the protective effect of oral GA+DL treatment. We were also able to show that the combination of glycolic acid and D-lactate in our specific formulation was well tolerated by the two ALS patients, even for up to 1.5 years treatment time in the case of the FUS patient.

These observations are in line with our recent study which showed that GA & DL restored axonal trafficking deficits of mitochondria and lysosome in FUS- and SOD1-ALS and rescued mitochondrial membrane potential as well as mitochondrial fragmentation (FUS-ALS) or elongation (SOD1-ALS) [29]. Of note, these effects were only seen in case of mitochondrial depolarization (i.e. FUS-ALS and SOD1-ALS) but not in TDP43-ALS, in which mitochondrial membrane potential is not disturbed [14]. This fits to previous data on PARK7 cell and C. elegans models, in which GA reversed the mitochondrial membrane potential and neuronal survival [41] which might indicate the transnosological therapeutical potential of GA/DL, specially in neurodegenerative diseases in which mitochondrial depolarization.

The reason for this transnosological therapeutical potential could be explained with the results from our own work and that from others showing that GA and DL have a multitarget effect as i) GA and DL can help maintaining calcium homeostasis by reducing calcium influx in the cell [23], ii) GA increases energy production [24] and iii) GA is metabolized to produced reduced glutathione [25] There are two distinguishing features in ALS-affected motor neurons. One is an increase in intracellular calcium due to several factors and the other is the alteration of mitochondrial function, which helps to increase intracellular calcium, reduces energy production, alters axonal transport and increases oxidative stress [42]. The reasons for the intracellular calcium increase are thought to be i) the excessive influx of Ca2+ ions due to high-frequency rhythmic activity and an increased expression of Ca2+-permeable AMPA receptors [43, 44], ii) these motor neurones exhibit low expression of Ca2+ buffering proteins like parvalbumin and calbindin [45]. Overexpression of these proteins has been shown to confer resistance to ALS-induced toxicity in motor neurones [46] iii) notably, ALS-associated mutations, including those in VAPB VAMP (vesicle-associated membrane protein-associated protein B and C gene), SIGMAR1 (sigma non-opioid intracellular receptor 1 gene) and SOD1 (superoxide dismutase 1 gene) can potentiate Ca2+ deregulation and heighten vulnerability to the effects of Ca2+ influx [47, 48] These findings emphasize the inherent susceptibility of motor neurones to intracellular Ca2+ overload as a significant risk factor for degeneration [49] and iv) mitochondria play a pivotal role in regulating intracellular Ca2+ levels through the mitochondrial uniporter, particularly crucial in motor neurones with low intrinsic cytosolic Ca2+ buffering capacity. In these vulnerable motor neurones, mitochondria are responsible for absorbing over 50% of intracellular Ca2+ increases, setting them apart from other cell types.

Consequently, the pronounced demand placed on specialized Ca2+ storage organelles make motor neurones even more susceptible to Ca2+ imbalances [50, 51], especially in ALS where mitochondria are frequently damaged [52]. Additionally, mitochondria impairment in ALS leads to decreased energy levels in the cell and increases in oxidative stress [42]. It would seem plausible, that GA and DL exert their therapeutic effect by reverting these alterations. Thus showing the potential of multitarget treatments in complex neurodegenerative diseases as ALS.

These preliminary data would suggest the convenience to perform more detailed studies searching for an appropriate biomarker and analyzing the feasibility of this combined therapy in a clinical set (maybe with combined other therapies addressed to other targets) with a well designed double-blind randomized against placebo trial to confirm the potential usefulness of GA+ DL in ALS treatment.

## Authoŕs contribution list

FP-M designed and coordinated the study. FP-M, AC, YD and AH, designed the mouse experiments. FP-M, FGD, GG and ALM designed the Drosophila experiments. FP-M, KB, JL and MD designed the protocol for the experimental treatment in ALS patients and obtained the permission from the ethical committee. AC and YD performed the mouse experiments. AJZ and GG performed the experiments in Drosophila. SS, CP, GR and RG monitored and controlled ALS patients. FP-M, ALM, AH and AC wrote the manuscript. All other authors critically revised and corrected the manuscript.

## Data Availability

All data produced in the present study are available upon reasonable request to the authors

## Acknowledgements

We would like to thank Olaf den Hollander from Corbion Purac and Amin Zrelli from Galactic for donating high purity D-Lactic acid to produce the drugs for the experimental treatment in patients. This work was funded by the Deutsche Forschungsgemeinschaft (DF, erman Research Foundation) under ermany’s Excellence Strategy within the framework of the Munich Cluster for Systems Neurology (EXC 2145 SyNergy – ID 390857198) and by the German Ministry for Economy and Energy with an EXIST-Forschungstransfer Grant (GLYMIPRO-FKZ03EFLBY173). AH is supported by the Hermann and Lilly Schilling Stiftung für medizinische Forschung im Stifterverband.

## Competing Interests

FP-M has a patent on the use of glycolic acid and D-lactate in ALS. He is also the founder of Neurevo GmbH and has obtained consulting fees from STADA and Bial. AT has obtained consulting fees from Roche and Astellas. All other authors declare no competing interests.

## Notes

### Funding Statement

This work was funded by the Deutsche Forschungsgemeinschaft (DFG, German Research Foundation) under Germany Excellence Strategy within the framework of the Munich Cluster for Systems Neurology (EXC 2145 SyNergy ID 390857198) and by the German Ministry for Economy and Energy with an EXIST Forschungstransfer Grant (GLYMIPRO FKZ03EFLBY173). AH is supported by the Hermann and Lilly Schilling Stiftung fuer medizinische Forschung im Stifterverband.

### Author Declarations

All proceedings described below were presented at the ethic committees of the University Hospital of the LMU (Reg. Nr.: 22-0366UE) and the Carl Gustav Carus University Hospital (Reg. Nr.: EK49022016) who showed no concerns regarding the design of the experimental treatment and publication of the results. Glycolic acid, D-Lactic acid and L-Alanine Ph.Eur. were obtained by the LMU hospital pharmacy. The quality control of Glycolic acid and D-Lactic acid was performed using the methods described in the Deutscher Arzneimittelcodex (German Drug Codex/NRF), L-Alanine Ph.Eur. was tested using the corresponding European Pharmacopoeia monograph.

