## Supplementary Figures for "The combination of glycolic acid and D-lactate delays disease progression in SOD-1 ALS mice, partially rescues lethality in iTBPH^pkk(108354)^ Drosophila and shows promising results in experimental treatments in two ALS patients"

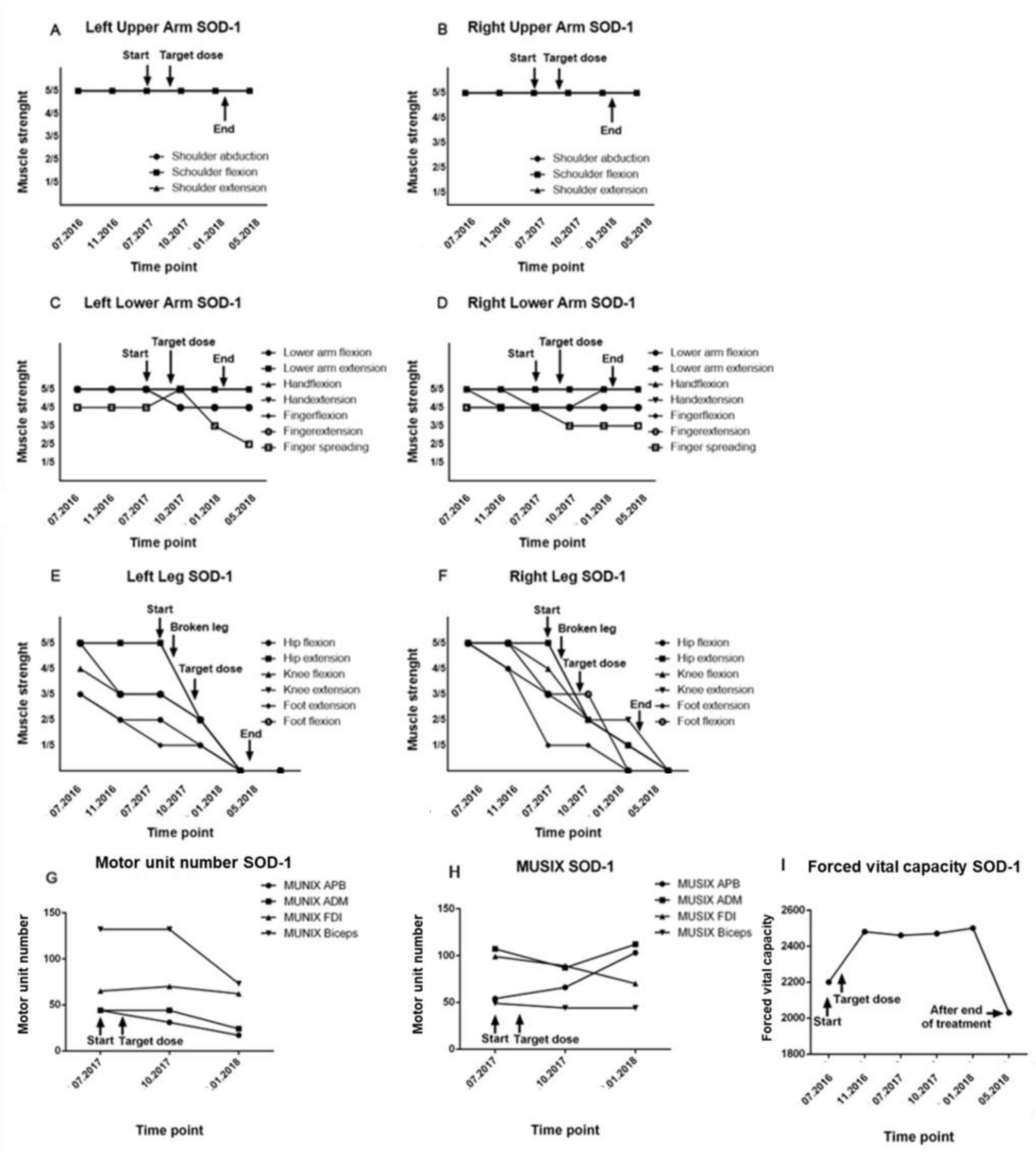


**Figure S1:. Disease progression SOD-1 ALS patient during treatment. (A)** Using MRC scale repetitive assessment of muscle strength of left upper arm (A), right upper arm (B), left lower arm (C), right lower arm (D), left leg (E), right leg (F). (G,H) Time course of the remaining number of motoneurons measured with Motor Unit Number Index (MUNIX) of hand and arm muscles. (I) Time course of forced vital capacity (FVC) measured with spirometry.

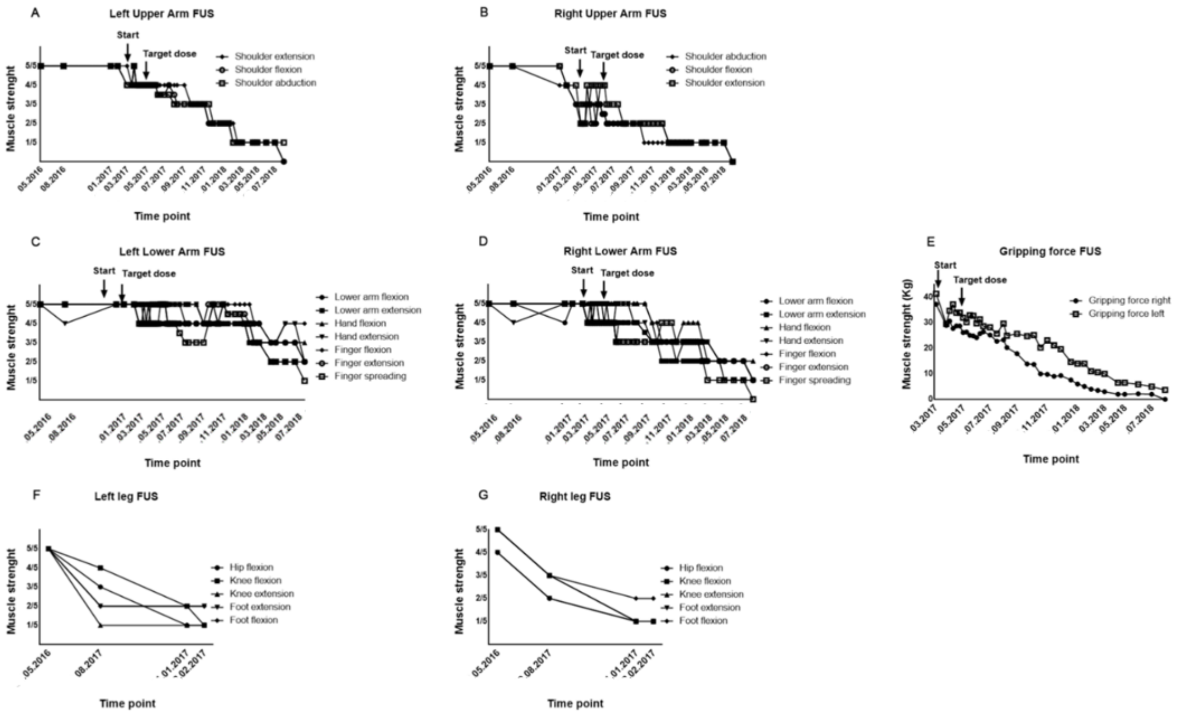


**Figure S2: Disease progression FUS ALS patient.** A-E show the disease progression as loss of strength in the different limbs. A and B show the strength in kg, as recorded using a digital dynamometer, C-E show the loss of strength based on clinical observations using the MRC-Scale. F and G show the loss of strength in the legs based on the examinations performed by the colleagues before treatment and described in previous medical reports. As it can be noticed, the FUS patient had lost the strength in the legs within 6 months without treatment before immediately starting to lose the strength in the upper limbs. This patient lost the strength in the upper limbs under the experimental treatment within 1,5 years.
